# Development, validation, and evaluation of prediction models to identify individuals at high risk of lung cancer for screening in the English primary care population using the QResearch^®^ database: research protocol and statistical analysis plan

**DOI:** 10.1101/2022.01.07.22268789

**Authors:** Weiqi Liao, Judith Burchardt, Carol Coupland, Fergus Gleeson, Julia Hippisley-Cox, DART initiative (WP6)

## Abstract

**Background and research aim:** Lung cancer is a research priority in the UK. Early diagnosis of lung cancer can improve patients’ survival outcomes. The DART-QResearch project is part of a larger academic-industrial collaborative initiative, using big data and artificial intelligence to improve patient outcomes with thoracic diseases. There are two general research aims in the DART-QResearch project: (1) to understand the natural history of lung cancer, (2) to develop, validate, and evaluate risk prediction models to select patients at high risk for lung cancer screening.

**Methods:** This population-based cohort study uses the QResearch® database (version 45) and includes patients aged between 25 and 84 years old and without a diagnosis of lung cancer at cohort entry (study period: 1 January 2005 to 31 December 2020). The team conducted a literature review (with additional clinical input) to inform the inclusion of variables for data extraction from the QResearch database. The following statistical techniques will be used for different research objectives, including descriptive statistics, multi-level modelling, multiple imputation for missing data, fractional polynomials to explore non-linear relationships between continuous variables and the outcome, and Cox regression for the prediction model. We will update our QCancer (lung, 10-year risk) algorithm, and compare it with the other two mainstream models (LLP and PLCOM2012) for lung cancer screening using the same dataset. We will evaluate the discrimination, calibration, and clinical usefulness of the prediction models, and recommend the best one for lung cancer screening for the English primary care population.

**Discussion:** The DART-QResearch project focuses on both symptomatic presentation and asymptomatic patients in the lung cancer care pathway. A better understanding of the patterns, trajectories, and phenotypes of symptomatic presentation may help GPs consider lung cancer earlier. Screening asymptomatic patients at high risk is another route to achieve earlier diagnosis of lung cancer. The strengths of this study include using large-scale representative population-based clinical data, robust methodology, and a transparent research process. This project has great potential to contribute to the national cancer strategic plan and yields substantial public and societal benefits through earlier diagnosis of lung cancer.

## Introduction

Lung cancer is a research priority in the UK. According to the most recent statistics from Cancer Research UK, lung cancer is the third most common cancer in incidence (after breast and prostate cancers), and the most common cause of cancer death in the UK. Incident lung cancer cases accounted for 13% of all new cancer cases, but lung cancer deaths accounted for 21% of all cancer deaths in 2017, more than twice of the second highest cancer mortality (bowel cancer, 10%).

Compared with other cancers, lung cancer survival is poor. Only 40.6% of patients survived one year or longer (2013-2017), and the 5-year and 10-year survival rates were 16.2% and 9.5%, respectively [1]. Early diagnosis of lung cancer could increase patients’ chances of receiving potentially curative treatments, and improve the poor lung cancer survival in the UK [2, 3].

Low-dose computerised tomography (LDCT) has been recommended for lung cancer screening by the United States Preventive Services Task Force (USPSTF) since 2013, for people between 55 and 80 years old, who have a history of heavy smoking and still smoke or quit smoking within the past 15 years [4]. The USPSTF updated their recommendation in 2021 by reducing the age threshold to 50 years and smoking exposure to 20 pack-year [5]. However, lung cancer screening using LDCT is still not a routine service in the UK at the moment. The NHS launched a new service from autumn 2019, the Targeted Lung Health Check (TLHC), for ever-smokers between 55 and 75 years old registered with a GP. The TLHC programme plans to deliver the service to approximately 600,000 eligible participants in 14 Clinical Commissioning Groups (CCGs) in England over four years (2020-2023) [6]. Patients outside those 14 CCGs may not be able to access this service, which could be a potential health equality issue of care access. Therefore, a population-based study that focuses on the development, validation, and comparison of prediction models for personalised lung cancer risk for the English population and the associated cost-effectiveness analysis may provide timely evidence for the UK National Screening Committee (UK NSC) to expedite decision making for lung cancer screening programmes in the four UK countries. Such health policy may help shift the diagnosis of lung cancer towards earlier stages, which can lead to better survival outcomes.

The DART project (full project title: The Integration and Analysis of Data using Artificial Intelligence to Improve Patient Outcomes with Thoracic Diseases) is an academic-industrial collaborative initiative funded by Innovate UK (UK Research and Innovation), led by the University of Oxford, and working closely with the TLHC programme. There are nine work packages (WP) for the whole project. Work package 6 (primary care, population health, and health economics) aims to identify potential opportunities for improved diagnostic and cost-effectiveness for lung cancer screening in the UK population. The statistical (risk prediction) models can measure and assess the effects of cancer risk across different timeframes, for example, short-term (1-year), medium-term (5-year), and long-term (10-year risk of developing lung cancer) and predict the likely impact of using different thresholds of lung cancer risk for LDCT scan at the population level. The health economic (cost-effectiveness) analysis can identify ineffective lung cancer screening strategies so that they can be refined or avoided.

### Research objectives of the DART-QResearch project

This research protocol covers the DART-QResearch part (WP6, primary care and population health). The objectives for this project are to:

1. Undertake a literature review to identify existing lung cancer prediction models and critically appraise these prediction models using the PROBAST tool [7];
2. Determine the current epidemiology for the natural history of lung cancer from first presentation, investigation, referral, diagnosis, treatment, and survival using data from the QResearch database, and examine how the natural history of lung cancer varies by age, sex, ethnicity, socioeconomic deprivation, smoking status, geographical regions and over time;
3. Identify and quantify the risk factors for lung cancer based on the analysis of electronic health records (EHRs) and compare the findings with the literature;
4. Update and validate the existing QCancer (lung) algorithm using more recent data linked to HES, death and cancer registries;
5. Compare the updated QCancer (lung) model with the other risk prediction models identified from the literature, and select and recommend the best model for population-based lung cancer screening.

## Study design and methods

### Data source – the QResearch® database

Routinely collected electronic health records (EHRs) linked to the QResearch database (version 45) will be the main data source for this project. QResearch is a large consolidated database with anonymised EHRs of over 35 million patients from 1800+ general practices using the Egton Medical Information Systems (EMIS) spread across England. The database includes patients who are currently registered with practices as well as historical patients who may have left or died. Historical records date back to 1989 with linked data on all practices since 1998. Patients’ primary care records are linked with other national datasets, such as the Hospital Episode Statistics (HES, secondary care data, including inpatient, outpatient, accident and emergency (A&E), and critical care), death registration (up to 15 causes of death) from the Office for National Statistics (ONS), and cancer registration data from Public Health England (PHE).

### Data preparation

The TRIPOD guideline [8] recommends seeking external evidence and critical consideration of relevant literature for selecting variables in the prediction model. The team conducted a rapid literature review (including the NICE guidelines) and had clinical input to inform the inclusion of variables and prepare the code lists to extract data from the QResearch database. We prepared Read/SNOMED-CT code lists to extract events from GP records, ICD-10 code lists for diagnosed diseases in the HES, cancer registry, and death records, and OPCS code lists for interventions and procedures conducted in NHS hospitals. This has been done at the study design phase, before submitting the research proposal to get approval from the QResearch Scientific Committee. The variables were included as broadly and comprehensively as possible for data extraction. Table 1 summarises the variables requested for the DART-QResearch project.

**Table 1.**
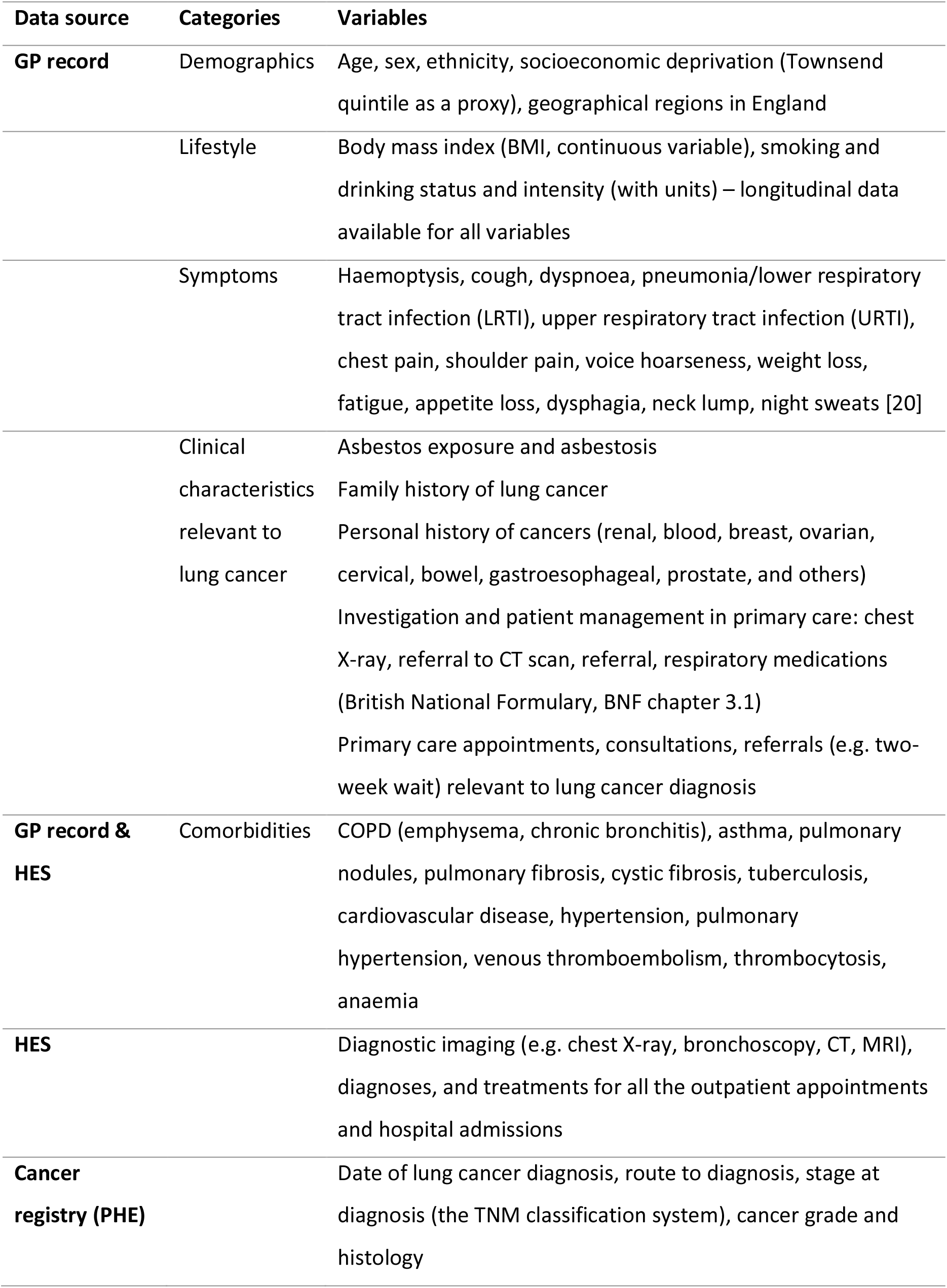
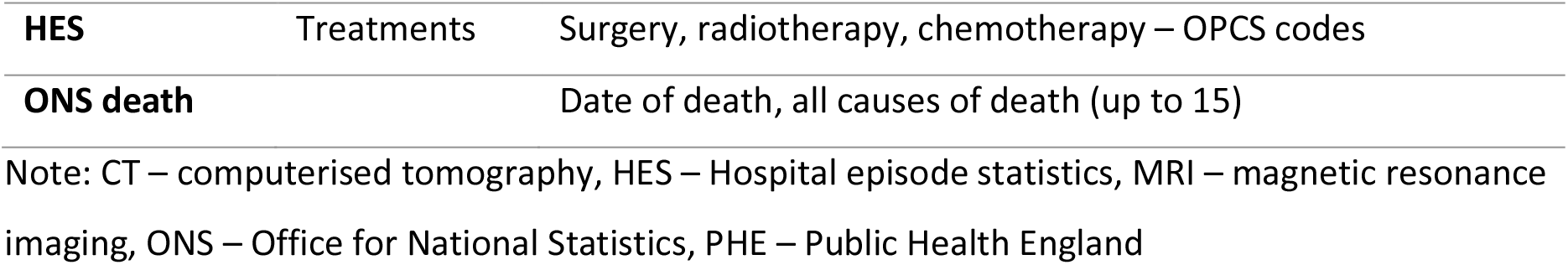
Variables extracted from the QResearch database for this project

### Study design, setting, and population

This is a population-based retrospective cohort study of the English primary care population. The study period is from 1 January 2005 to 31 December 2020. We will use similar inclusion and exclusion criteria as those in the previous studies [9-12] to develop and validate the QCancer models. The study population will be adult patients aged between 25 and 84 years old and without a diagnosis of lung cancer before entering the cohort. The patients need to be registered in the general practices for at least 12 months, and these practices have contributed to the QResearch database for a minimum of 12 months before the cohort entry date. This is to ensure complete data before cohort entry.

The age range is wider than the TLHC programme (55-75 years) [6], the USPSTF recommendation for lung cancer screening (55-80 years) [4], the Liverpool lung project (LLPv2 and LLPv3) prediction model (the English population, 55-75 years) [13, 14], and the PLCOM2012 prediction model (the American population, 55-74 years, where PLCO stands for the Prostate, Lung, Colorectal and Ovarian cancer screening trial) [15]. This is because we intend to compare our model with the other mainstream models for lung cancer screening. In addition, QResearch has rich clinical data on comorbidities, personal history, and family history, which allows us to assess the risk of patients aged under 50 years for early-onset lung cancer. Therefore, this broad age range covers the majority of patients (inclusivity) and allows more flexibility to produce research evidence on which populations are more likely to benefit from active surveillance and screening for early diagnosis of lung cancer. For patients older than 85 years, cost-effectiveness and over-diagnosis would be the key concerns, and the benefit of screening may be marginal.

### Identification of cases

Incident lung cancer cases during 2005-2018 (the most recent available data from the linked cancer registry) will be identified from the four linked data sources, and followed up to 31 March 2020. Data for treatments and outcomes (e.g. death, left cohort, still alive) for the cancer cases are available in the follow-up period in the HES and ONS datasets. Patients with previous or secondary diagnosis (metastasis) of lung cancer.

### Pathways to lung cancer diagnosis

Figure 1 illustrates the milestone events and different intervals in the cancer care pathway from the first symptom to the start of treatment [16]. The following key concepts and intervals defined in the Aarhus statement [17] are of interest in research objective 2. Referral, diagnostic, and treatment intervals will be explored, as NHS England set national waiting time targets for these intervals in the cancer care pathway (summarised in Table 2).

- **Date of the first presentation**: the date that the patient presented in general practice with signs or symptoms probably due to cancer within 1 year before diagnosis [18, 19];
- **Date of referral**: the date that GP sent the referral letter;
- **Date of diagnosis**: the earliest date of lung cancer diagnosis recorded in primary care, secondary care records, cancer or death registry in the QResearch database;
- **Diagnostic interval**: the duration from patient’s first presentation in primary care within the 12 months before diagnosis to the date of a confirmed diagnosis of lung cancer (examples of empirical studies [18, 19]);
- **Treatment interval**: the duration between confirmed cancer diagnosis and the start of cancer treatment recorded in the HES dataset.

**Figure 1.**
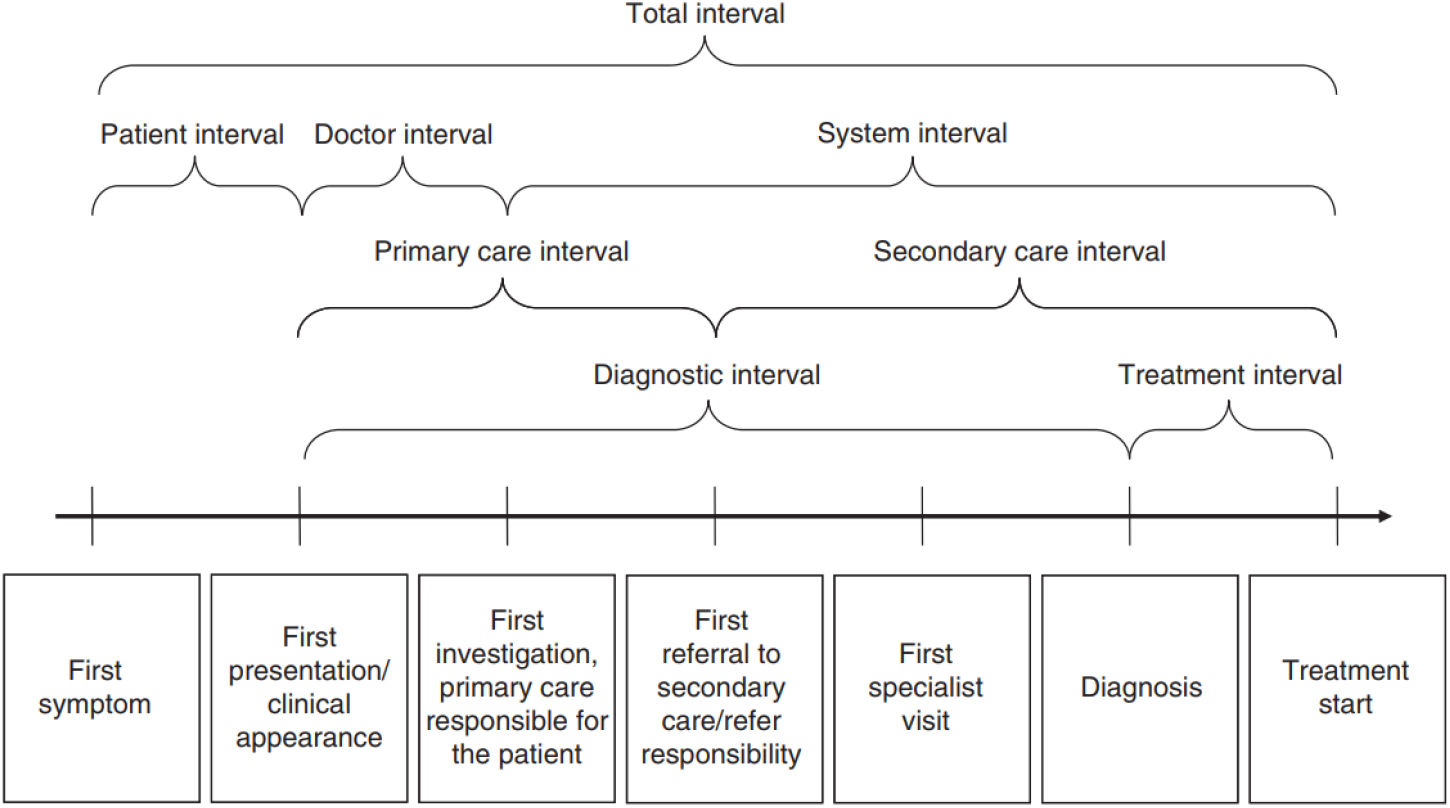
The conceptual model for the cancer care pathway. The conceptual model for cancer care pathway showing the milestone events and intervals from the first symptom to the start of treatment. Reference: [16]

**Table 2.**
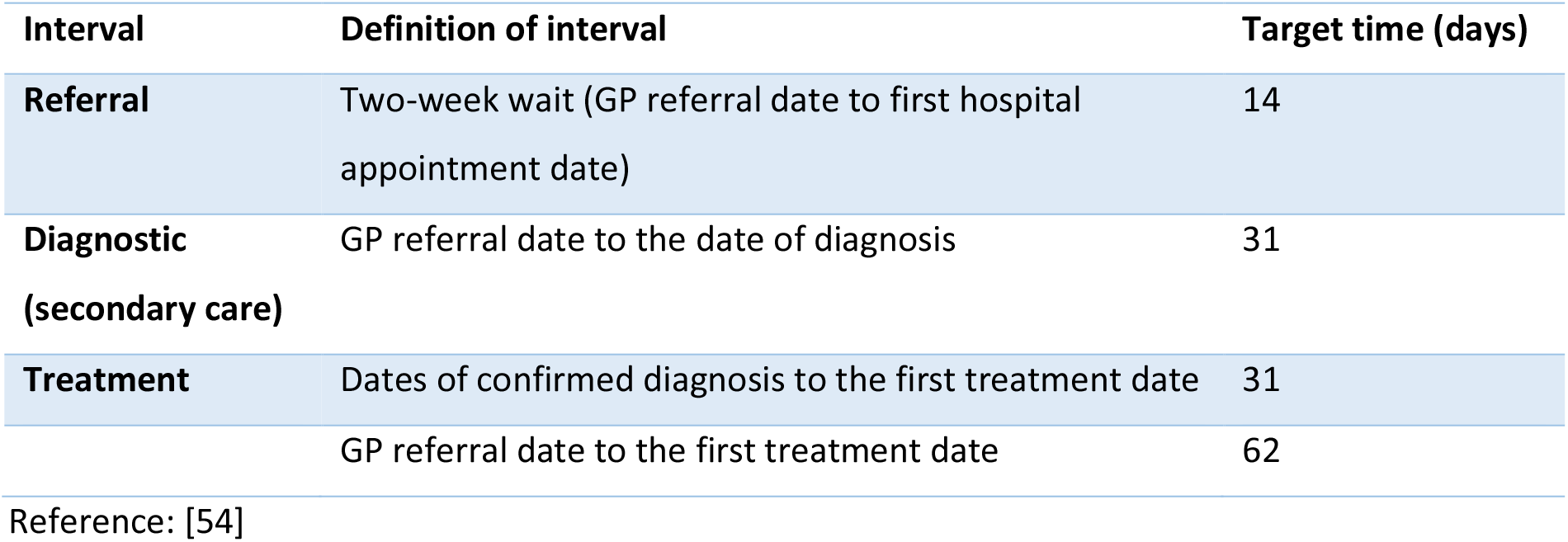
National waiting time targets for cancer in the NHS

### Outcomes for the prediction model

The primary outcome for the prediction model (research objectives 3-5) is the incident diagnosis of lung cancer. Code lists are published https://www.qresearch.org/qcode-group-library/. We will use the earliest date on any of the four linked databases as the date of lung cancer diagnosis. The secondary outcomes are stage at diagnosis (likely to convert into a binary variable, i.e. early vs late stage) and death due to lung cancer.

### Ethical approval of the project

The DART-QResearch project has obtained approval from the QResearch Scientific Committee on 8 March 2021. QResearch is a Research Ethics Approved Research Database, confirmed from the East Midlands – Derby Research Ethics Committee (Research ethics reference: 18/EM/0400, project reference: OX37 DART). A dedicated webpage for this project has been created on the QResearch website https://www.qresearch.org/research/approved-research-programs-and-projects/the-integration-and-analysis-of-data-using-artificial-intelligence-to-improve-patient-outcomes-with-thoracic-diseases-dart/. The lay summary of this project for the public is available on this webpage.

### Patient and Public Involvement and Engagement (PPIE)

We are very fortunate to have regular lay member representatives from the Roy Castle Lung Cancer Foundation involved at the beginning of this project, to review our lay summary and provide feedback as part of our ethical approval. The Roy Castle Lung Cancer Foundation is a charity dedicated to helping people affected by lung cancer in the UK. It is a PPIE partner for the whole DART project (9 work packages). In addition, as the study goes on, we will engage more patient representatives and involve relevant stakeholders when we disseminate our study findings and ask for their comments. Constructive feedback from wider NHS service user groups and academic audiences is welcome.

## Statistical analysis plan

### Natural history of lung cancer (research objective 2)

#### Symptomatic presentation for patients diagnosed with lung cancer

A previous systematic review [20] identified symptoms significantly associated with lung cancer. We will include those symptoms in our analysis. We will describe and compare patients’ symptomatic presentation in 3, 6 and 12 months before the diagnosis of lung cancer. In addition, patients may present to their GP with several different symptoms. The most common symptom combinations will be summarised. These findings may help GPs pick up symptoms and consider lung cancer earlier, and manage patients accordingly in the disease trajectory. Some patients may not have any symptoms recorded in primary care EHRs, as they may present in A&E (in emergency presentation route). Sequence analysis [21] will be used to construct **symptom trajectories** leading to the diagnosis of lung cancer for patients with symptomatic presentation and calculate the dissimilarity between sequences. Cluster analysis (agglomerative hierarchical clustering, Wald’s method) [22] will be used to group similar sequences based on the dissimilarity (distance) between symptom trajectories. The final results will be different **symptom phenotypes** (clusters) of symptom trajectories.

#### Referral, diagnostic, and treatment timeliness and the influencing factors

Descriptive statistics will be used to describe the sociodemographic and clinical characteristics (e.g. comorbidity, cancer stage, grade, histology) of the study population, using means and standard deviations, medians and interquartile ranges (IQR), and proportions as appropriate. The referral, diagnostic, and treatment intervals between the milestone events in the natural history of lung cancer will be calculated using day as a unit. The distribution of each interval variable will be checked. Line charts will be made to show the temporal changes in the diagnostic and treatment intervals, routes to diagnosis, stage at diagnosis, and treatment (surgery, radiotherapy, chemotherapy) of lung cancer cases from 2005 to 2020.

Parametric (e.g. ANOVA) and non-parametric statistical tests (e.g. chi-square test, Kruskal-Wallis test, where appropriate) will be used to investigate whether there are any significant differences in the diagnostic and treatment intervals of lung cancer by age (continuous variable), sex (binary variable), ethnicity, socioeconomic deprivation (Townsend quintile as a proxy), smoking status, geographical region (categorical variables). The association between the number of symptoms recorded in primary care EHRs, the number of visits to general practice, diagnostic interval, and cancer stage will be explored. Multi-level modelling (2 level random intercept model) will be used to explore the practice effect in the diagnostic interval (continuous variable), where level 1 is individual patient and level 2 is general practice (random effect, patients clustered in practices). Patients’ sociodemographic, clinical characteristics, and relevant interaction terms (e.g. age, sex, socioeconomic deprivation) will be considered and included in the model. Such analyses aim to explore:

1. whether certain patient characteristics would influence/increase diagnostic interval (e.g. older male patients in lower SES with long-term smoking habit);
2. whether certain clinical features (e.g. the number of potential lung cancer symptoms, cardiorespiratory comorbidities such as COPD, asthma, hypertension, etc.) and indicators in primary care services (the number of primary care visits 1 year before diagnosis) are associated with the diagnostic interval;
3. whether there is a practice effect in the diagnostic interval: whether certain practices performed better than others (i.e. patients in some practices consistently had shorter diagnostic intervals, or diagnosed at early stages).

### Methodology of development, validation, and comparison of prediction models (research objectives 3-5)

#### Sample size considerations

Sample size calculations for a risk prediction model will ensure precise estimation of the model parameters whilst minimising potential overfitting. We used the criteria by Riley et al. [23] and the ‘pmsampsize’ package in R to calculate the minimum required sample size for developing a clinical prediction model. The parameters for sample size estimation for time-to-event outcome were set or assumed as follows. The previous QCancer prognostic models [12] have around 30 predictors, we assume 50 predictors in the updated models to allow more flexibility. The median duration from cohort entry to the incident diagnosis of lung cancer is about 6 years, and the maximum predictive period is up to 10 years (QResearch has linked data on all practices since 1998, and the study period is 2005-2020). According to statistics of lung cancer incidence from Cancer Research UK [24], the age-standardised incidence rate (event rate) of lung cancer in the UK during 2016-2018 was 90.6 (95% CI: 89.9-91.2) per 100,000 population in men and 70.1 (95% CI: 69.6-70.7) in women. A conservative 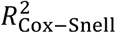 (15% of the maximum 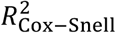) was used as recommended [23]. Based on the above parameters, the minimum sample size required for developing a new model is 42,607 for men and 59,750 for women. Hence, a minimum total sample size of about 102,500 men and women for model development is needed.

With over 18 million patients in the open cohort and an estimated 84,000 incident cases of lung cancer during 2005-2018 in the QResearch database, there is sufficient data for the development and validation datasets. We will use all the eligible patients in the database to maximise the power.

#### Exploration of non-linear relationships

Before imputation, a complete-case analysis will be fitted using a Cox model containing only the continuous variables (e.g. age, BMI) within the development dataset to derive the fractional polynomial terms (up to two polynomial terms) [25] for non-linear relationships. Separate models will be fitted for men and women.

#### Handling (missing) data

For comorbidities, personal history and family history, the absence of information in the EHRs is assumed that the patient did not have the health conditions or family history of those conditions. There may be missing data in some other variables, as they may not have been collected and recorded in the EHRs, particularly in the early years. We will use multiple imputation with chained equations (MICE) to replace missing values for ethnicity, Townsend quintile, BMI, smoking status, alcohol intake, and stage at diagnosis with the assumption of data missing at random (MAR) [26-29]. Five imputations will be conducted, as this has relatively high efficiency [8] and is a pragmatic approach accounting for the amount of data and the capacity of the available computing power in the software and the server. Rubin’s rules will be used to combine the parameter estimates for the model across the imputed datasets [30].

#### Model development

Three-quarters of general practices will be randomly selected for model development, and the remaining quarter of general practices will be for validation (internal validation approach). Separate models will be developed and validated for men and women, as the coefficients of predictors may be different between sexes, also making the computing power more feasible considering the sample size.

We will use similar established analytical strategies to develop and validate the risk prediction models in this study that were used in the previous QResearch studies [12, 31-35]. Cox proportional hazards model will be used as the main method to develop the risk prediction models, to identify significant patient and clinical characteristics for incident diagnosis of primary lung cancer and estimate the hazard ratios, using robust variance estimates to allow for clustering of patients within general practices, also accounting for censoring in the cohort. The assumption of proportional hazards for Cox regression will be checked. The risk period of interest is from the date of entry to the study cohort to the date of incident diagnosis of lung cancer. Patients who do not develop lung cancer will be censored on the exit date of the cohort (i.e. 31 March 2020). The main analyses will be multivariable analyses after multiple imputation for missing values, including various predictors and interaction terms. Complete case analysis will be conducted as additional sensitivity analysis. The model can be used to derive individualised risk estimates of developing lung cancer for each year of follow-up, for up to 10 years.

#### Variable selection and considerations

We will fit the models by including all the variables initially, and then retain those having a hazard ratio (HR) <0.90 or >1.10 (clinical significance) for binary and categorical variables and at the statistical significance level of 0.01 (two-tailed). For some less common variables, such as previous diagnoses of other cancers, family history of cancer, exposure to asbestos or asbestosis, we will retain the variables at the significance level of 0.05, since these events are rare and there may be small numbers for these variables. According to the TRIPOD guideline [8], the backward elimination approach in multivariable modelling is preferred. To simplify the models, we will focus on the most common health conditions, and combine similar variables with comparable HR where appropriate. If some variables do not have enough events to obtain point estimates and standard errors, we will combine some of these if clinically similar in nature. Otherwise, we will exclude them from the models.

#### Risk equations

The regression coefficients for each variable in the final model will be used as weights. From which, we will derive the risk equations by combining with the baseline survival function evaluated for each year of follow-up, up to a maximum of 10 years [36]. We will use the risk equations to estimate the absolute risk, with a specific focus on 5-year, 6-year, and 10-year risk, as we are interested in comparing our model with other validated prediction models for lung cancer screening, such as the LLP and PLCOM2012 models (separate subsection below). The baseline survival function will be estimated based on values of zero for centred continuous variables and all binary and categorical predictors.

#### Model validation – evaluate the model performance

An imputation model (MICE) will be fitted for missing values in the validation dataset for five imputations (same as in the deviation dataset) using the methods described in the earlier subsection. We will apply the risk equations for men and women derived from the previous step to the validation data and calculate the measures of model performance.

As in previous studies [37], we will calculate the R^2^ [38], the D statistic [39], the Brier score [40], and Harrell’s C statistics [41] at 5, 6, and 10 years and combine these across the imputed datasets using Rubin’s rules. R^2^ is the explained variation, where a higher value indicates a greater proportion of variation in survival time is explained by the model [38]. The D statistic is a measure of discrimination, which quantifies the separation in survival between patients with different levels of predicted risk, where higher values indicate better discrimination [39]. The Brier score is an aggregate measure of disagreement (the average squared error difference) between the observed and the predicted outcomes [40]. The Harrell’s C statistic [41] is a measure of discrimination (separation) that quantifies the extent to which those with earlier events have higher risk scores. Higher values of Harrell’s C indicate better performance of the model for predicting the relevant outcome. A value of 1 indicates that the model has perfect discrimination. A value of 0.5 indicates that the model discrimination is no better than chance. The 95% confidence intervals for the performance statistics will be calculated to allow comparisons with alternative models for the same outcome and across different subgroups [42].

We will assess the calibration of the risk scores by comparing the mean predicted risks at 5, 6, and 10 years with the observed risks by categories of the predicted risks (e.g. by decile or twentieth), which will be presented in calibration plot. The observed risks for men and women will be obtained by using the Kaplan-Meier estimates. We will also evaluate these performance measures in five pre-specified age groups (25-49; 50-59; 60-69; 70-79; 80+).

#### Updating the QCancer (10-year risk) model and comparing it with other mainstream prediction models for lung cancer screening

We will update the existing QCancer (lung, 10-year risk) model, as the QResearch database has been expanding rapidly over the last 5-10 years and now more data are available, especially for important variables such as stage at diagnosis, cancer histology and grade, which were not available when the QCancer (lung, 10-year risk) models were initially developed. We will also compare our model with other widely used algorithms to select patients for lung cancer screening using LDCT, such as the LLP models (v2 and v3) for 5-year risk [13], the PLCO models (both the original M2012 model for ever-smokers [15] and the updated M2014 model including non-smokers [43]) for 6-year risk. We will calculate measures of performance described above to compare the algorithms in different patient subgroups (e.g. patients in different age groups). Decision curve analysis [44] will be used to evaluate and compare the net benefit of the prediction models (clinical usefulness). We will compare different models with the same validation dataset, evaluate model performance, and discuss the strengths and limitations of each model for lung cancer screening, especially for the English primary care population. We will follow the recommendations from the TRIPOD guideline [8] to report the multivariable prognostic model.

#### Risk stratification

Risk stratification allows patients with a high predicted risk to be identified electronically from primary care records for tailored advice, active monitoring of disease progression, and lung cancer screening. We will examine the distribution of the predicted risks and calculate a series of centile values in the model. For each centile threshold, we will calculate the sensitivity and specificity of the risk scores. The currently accepted threshold for classifying high risk is 3% for the QCancer models in the NICE guideline [45]. The NHS England Targeted Lung Health Check programme uses either a 5-year risk threshold of 2.5% in the LLPv2 model and/or a 6-year risk threshold of 1.51% in the PLCOM2012 model as eligibility criteria [46].

#### Dissemination and implementation plan of the prediction model

The risk prediction algorithm will be published in a peer-reviewed journal and presented at academic conferences. A web-based program could make the updated risk algorithm publicly available in a similar way to the QCancer tool (https://www.qcancer.org/), subject to funding and Medicines and Healthcare Products Regulatory Agency (MHRA) medical device compliance. It will also be possible to implement the risk algorithm in the EHR systems, using existing data to calculate individual risks for the primary care population. These implementation intentions will be subject to the terms and conditions of QResearch, the University of Oxford, the Innovate UK grant, and the agreement of all parties. The implementation of the prediction algorithm will be covered by another protocol, which is out of the scope of this research protocol.

### Summary: relevant guidelines used in this study

- NICE guideline NG12 (Suspected cancer: recognition and referral) [45]
- The Aarhus statement (recommendations for research in early cancer diagnosis) [17]
- REST (Reporting studies on time to diagnosis) [47]
- The STROBE statement (reporting guideline for observational studies) [48]
- The TRIPOD statement (reporting guideline for a multivariable diagnostic or prognostic prediction model) [8, 49]
- The PROBAST tool (to assess the risk of bias and applicability of prediction model) [7]

## Discussion

### Methodological strengths and limitations of this study

#### Strengths

The key strengths of this population-based study include prospective recording of outcomes, good ascertainment of lung cancer cases through multiple record linkage, and a large sample size from an established and validated database which has been used to develop many risk prediction tools, such as QFracture [32], QRisk3 [34], QDiabetes [35]. A wealth of data are available for identifying risk factors and developing the prediction model. The UK primary care records have high levels of accuracy and completeness of clinical diagnoses and prescribed medications. This study has good face validity, as primary care has coverage of almost the entire population in the UK, and this study is conducted in the same setting where most patients are clinically assessed, managed, and followed up. Prediction models developed using primary care EHRs are likely to generalise to the wider English population. In addition, we intend to externally validate the LLP and PLCO models using English primary care data and compare the QCancer prediction model with the LLP and PLCO models using the same dataset. The findings could be used to inform which algorithm is most useful to select eligible English primary care population for lung cancer screening. This study also minimises the most common biases in epidemiological studies, such as selection bias, recall bias, and respondent bias. We also use relevant guidelines, statements, and recommendations for the research process and statistical analyses in this project. We publish this research protocol to promote transparent and reproducible research. All of these are the strengths of this project.

#### Potential limitations

Limitations of this project may include potential information bias and missing data. Some diagnoses in the primary care records lack formal adjudication. Based on our experiences of using primary care data, some lifestyle factors such as BMI, smoking and alcohol drinking status may not always track the true values in real-time. In addition, the recording of family history of cancer in primary care records may be sparse. As to the cancer registry data, the cancer stage is not complete (about 30% missing in 2017, even more before 2010), which limits further exploration of developing models to predict early versus late diagnosis of lung cancer. However, we may overcome this limitation by imputing cancer stage, as we have rich clinical data, treatments, and survival outcomes linked to the QResearch database, which can be used in the imputation model.

Due to the available resources, we will validate the developed model using data from the same database (QResearch uses data from the EMIS, which is the computer system used by 55% of UK GP surgeries). Our study population is based in England and representative of the whole English primary care population. The models will need to be evaluated if used outside of England. A more stringent approach would be using data from different EHR systems, different data sources (e.g. CPRD, THIN), or other countries in the UK (external validation). However, some previous independent studies [50-52] have examined other risk equations developed by the QResearch team and concluded that validation using external data showed similar levels of performance as the internal validation approach using the QResearch database, which is reassuring.

### Clinical implications for practice and future research

Lung cancer is the biggest cause of cancer death in the UK, and it is a research priority in this country. It cost an estimated £307 million in hospital care in 2010 [53], which is a huge burden to the NHS and society. Earlier diagnosis is crucial to reducing lung cancer mortality, care costs, and patient concerns. We hope to identify useful patterns from the natural history of lung cancer to enable GPs to recognise lung cancer earlier and better manage patients (Research objective 2). Developing risk-stratification models can help identify patients at high risk of developing lung cancer, and refer them to the TLHC programme or LDCT scan for early diagnosis, without unduly burdening the overstretched NHS (Research objectives 3-5). In addition, health economic analysis will provide new insights to maximise the cost-effectiveness of lung cancer screening (separate linked protocol). The potential impact includes earlier diagnosis and better survival outcomes for patients, reduced cost for the NHS, and a reduced disease burden for society.

## Data Availability

Due to the sensitive nature of anonymised patient level data (electronic health records), the study data are only accessible to the named researchers approved by the ethics committee.

## Declarations

### Authors’ contributions and consent for publication

FG and JH-C secured the funding. FG is the chief investigator of the DART project, and JH-C is the joint package lead and the guarantor of this project. JH-C and WL contributed to the study conceptualisation. WL specified the data, led on the ethical approval, and is the lead statistician for the DART-QResearch project. WL designed the statistical analysis plan and drafted the whole research protocol, with methodological input from JH-C and CC, clinical and contextual input from JH-C and JB. All authors read and commented on the earlier drafts, contributed to the revision of the manuscript, and approved the final version of the manuscript for publication.

## Acknowledgements

We thank the two lay members from the Roy Castle Lung Cancer Foundation reviewed our lay summary of the DART-QResearch Project for ethical approval and provided very helpful feedback.

This project involves data from patient-level information collected by the NHS, as part of the care and support for the patients. We acknowledge the contribution of the patients and general practices contributing to the EMIS (Egton Medical Information Systems) clinical computer system and the QResearch database, and the Universities of Nottingham and Oxford for the expertise in establishing, developing, and supporting the QResearch database. The Hospital Episode Statistics data used in this study are re-used with permission from NHS Digital, who retain the copyright of the data. The cancer registration data are supplied by Public Health England. The death registration data are provided by the Office for National Statistics. None of these organisations has been involved in any research process, including study design, data specification, statistical analysis, interpretation of results, preparing manuscripts, or the decision to publish.

## Authors’ information

Twitter:

@dartlunghealth (the DART project), @WLiao_Ox (Weiqi Liao), @JudithBurchardt (Judith Burchardt), @JuliaHCox (Julia Hippisley-Cox)

Project website: www.dartlunghealth.co.uk.

